# Food Insecurity Increases in Brazil from 2004 to 2018: Analysis of national surveys

**DOI:** 10.1101/2020.10.22.20217224

**Authors:** Rosana Salles-Costa, Aline Alves Ferreira, Ruben Araujo de Mattos, Michael E. Reichenheim, Rafael Pérez-Escamilla, Ana Maria Segall-Corrêa

**Affiliations:** Institute of Nutrition Josué de Castro, Federal University of Rio de Janeiro; Institute of Social Medicine, Rio de Janeiro State University, Rio de Janeiro, RJ, Brazil; Department of Social and Behavioral Sciences, Yale School of Public Health, New Haven, CT, USA; Food, Nutrition and Culture Program, Oswaldo Cruz Foundation, Brasília, DF, Brazil

**Keywords:** Food and Nutrition Security, Hunger, Time Series Studies, Health Surveys, Brazil

## Abstract

**Objective:** Describe secular changes in food security (FS) and severe food insecurity (FI) in Brazil.

**Design:** We analyzed four national surveys that assessed FI with the Brazilian Household Food Insecurity Measurement Scale (EBIA), and estimated the percentage changes of FS/FI levels between 2004 and 2013 (1st period) and between 2013 and 2018 (2nd period) by sociodemographic variables.

**Setting:** Data from the cross-sectional Brazilian National Households Sample Surveys (2004, 2009 and 2013) and Household Budget Survey (2018).

**Participants:** Nationally representative samples of household surveys (2004=112,530; 2009=120,910;2013=116,196; and 2018=57,920).

**Results:** The 1^st^ period was marked by a significant increase in FS (+18.9%) and by a reduction in severe FI (−53.6%). The 2^nd^ period showed a decrease in FS (−18.2%) and an increase of severe FI (+43.8%). The greater increase FS in the 1^st^ period was in the Northeast (+33.4%), among households with more than 7 residents (+40.8%), and in households where the reference person self-identified as black or mulatto (+27.6%). In the 2^nd^ period, the lower increase in severe FI was observed among households with children under 4 years old (+ 6.3%) and with members over 65 years old (+12.5%).

**Conclusions:** After a significant reduction in FS from 2004 to 2013, FS was strongly compromised from 2014 to 2018 likely as result of disruptions in access to foods in all regions, intensified by the sociodemographic inequality in Brazil. Hunger in Brazil has re-emerged as a national concern.

## INTRODUCTION

Although governments across the world have committed to ending hunger, food insecurity (FI) and malnutrition by 2030, the attainment of this key Sustainable Development Goal (SDG) remains off track with dire consequences for human and planetary health^(1)^. The United Nations Food and Agricultural Organization (FAO) defines FI as the condition that exists *“when people lack secure access to sufficient amounts of safe and nutritious food for normal growth and development and an active and healthy life”* ^(2)^. According to recent FAO (3) (FAO 2020), 690 million people are hungry, representing almost 9% of the global population. Furthermore, FAO reports that nearly one in ten people in the world are exposed to severe forms of FI, with a major concentration of severe FI in Africa, Southern Asia, and Latin America ^(3)^.

Starting in 2003, the Government of Brazil launched major social policies and programs to improve food and nutrition security (FNS). These included the Zero Hunger Strategy (*Estratégia Fome Zero*)^(4,5)^ and the conditional cash transfer program called *Programa Bolsa Família* ^(6,7)^. In addition, the Brazilian government introduced measures to regulate food prices to reduce the cost of the basic food basket and built food safety stocks ^(8)^. In addition, the Government strengthened the legal framework for FNS, with the participation of the National Food and Nutrition Security Council (CONSEA) in the policy process ^(9)^ (Perez-Escamilla, 2012). The FNS National System was established to ensure the human right to adequate food^(10)^. In 2010, access to food became a social right established by the Brazilian Constitution. In the same year, the National Policy for Food and Nutrition Security, PNSAN^(11)^, was established. According to scholars^(9)^, these initiatives and institutional developments were fundamental in enabling Brazil to leave the Hunger map in 2014. The strong political commitment followed by legislation was indeed decisive for the reduction of hunger and extreme poverty^(13)^, and the reduction in household FI, especially severe FI ^(14)^.

The documentation of changes over time in food security (FS) and different levels of FI in the population was possible due to the introduction, adaptation, and validation in Brazil of an experience-based food security scale, i.e. the Brazilian Food Insecurity Scale (EBIA for its acronym in Portuguese) ^(9,15-17)^. Experience-based scales are now being used in many countries, including Brazil^(18,19)^. Indeed, FAO now tracks SDG 2.2, with the Food Security Experience Scale (FIES)^(20)^.

The EBIA was developed based on an experience-based scale adapted from the US Household Food Security Survey Module (HFSSM), used in population surveys since the early 1990s^(21,22)^. The EBIA, whose validation began in 2004^(23)^ followed by additional refinements^(24,25)^, is based on the premise that FI is a phenomenon perceived and experienced by families at different levels of severity. The use of the EBIA has contributed information and strategic data for the management of policies, programs and actions directly related to the fight against hunger and poverty ^(9,26)^ and has been fundamental for the evaluation and monitoring of the dimensions related to the SDGs in the 2030 Agenda^(27)^ and a valuable tool for analyzing FNS governance in Brazil^(9)^.

In Brazil, the Brazilian Institute of Geography and Statistics (*Instituto Brasileiro de Geografia e Estatística – IBGE)* incorporated the EBIA in the *Brazilian National Household Sample Survey* (PNAD) from 2004, 2009 and 2013, allowing for the analysis of FI secular trends in the Brazilian population for almost a decade ^(28)^. In 2017, the IBGE included the EBIA in the module on living conditions of the Brazilian population Family Budget Survey (*Pesquisa de Orçamentos Familiares - POF*) to continue estimating the prevalence of household FS/FI in Brazilian, and enabling to analyze the FI data with respect to robust family expenses and personal food consumption indicators. The POF, like the PNAD, is representative at the national level as well as for urban and rural households by country macroregion. The first set of findings from the POF 2018 were released in September 2020^(29)^ making it timely to update the FI/FS secular trend analyses in Brazil.

The aim of this study was to assess trends and variations in FS and FI, especially severe FI, by key demographic and socioeconomic variables, in Brazil from 2004 to 2018.

## METHODS

### Samples

Brazil is a heterogeneous country territorially divided into five sociocultural and economically distinct macro regions (North, Northeast, Midwest, Southeast and South). The first two regions are formed by the least-developed municipalities, as shown by their low average household incomes, low levels of education and poor health outcomes compared with those of the South, Southeast and Midwest regions^(30)^. This report is based on data from the PNADs collected in 2004, 2009 and 2013 and the POF from 2018, all surveys carried out by the IBGE following best practices including strong data quality control procedures. Their main purpose is to generate indicators that are useful for the timely monitoring of the social and economic development of the country.

The PNADs employed a three-stage probabilistic cluster sampling design, with the selection of municipalities in the first stage, census tract in the second stage and households in the third stage. Regarding the POF, the sampling plan involved a stratified, two-stage probabilistic cluster sampling design, with the selection of census tracts as PSUs and households in the second stage. The selection of PSUs employed a probability proportional to size sampling scheme according to the number of private households per census tract. The total number of PSUs was determined according to the type of estimator used and the level of precision set for estimating the total data for the households, obtained from the 2010 Demographic Census data, also taking into account the number of households in each census sector. The final number of households in each survey were: PNAD 2004=112,530; PNAD 2009=120,910; PNAD 2013=116,196; and POF 2018=57,920. PNADs and POF 2018 analysis estimates were weighted, considering the sampling design, and adjustments needed to compensate for the nonresponse of the included units.

In both the PNAD and the POF, data were collected by trained interviewers meeting faceto-face with residents. In the POF, data collection was performed on consecutive days during the nine-day period using portable computers for registration and data entry. The database went through data quality control to assess the coherence of the information by trained technical staff. Further details of the sample design, the total number of PSUs interviewed by state, data quality control and the imputation of variables are described in the IBGE official report ^(31-34)^.

### Assessment of Household Food Insecurity

EBIA’s raw score was used to classify households into the following mutually exclusive FS/FI categories using recommended cut-off point for households with and without minors^(25)^: (i) FS (when the family/household has regular and permanent access to quality food, in sufficient quantity,); (ii) mild FI (presence of concern or uncertainty about access to food in the future; presence of inadequate food quality resulting from objectives that aim not to compromise the quantity of food); (iii) moderate FI (quantitative reduction of food among adults and/or disruption in eating patterns resulting from lack of food among adults); and (iv) severe FI (quantitative reduction of food also among those under 18 years of age, that is, disruption in eating patterns resulting from the lack of food among all residents; in this situation, hunger becomes a lived experience at home).

Since its first use in PNAD 2004, EBIA has undergone adaptations regarding the number of items used to evaluate FS/FI levels, maintaining the comparability between the prevalence estimated across surveys. The decisions to review EBIA was the result of a meeting of experts and its psychometric validity was confirmed with the Rasch model. hence. The decision to reduce the number of scale items from 16 to 14, did not affect the scale’s internal validity^(25)^. Additionally, the cutoff points used to classify the levels of FS/FI have been equivalents since 2004. The EBIA consists of 14 dichotomous questions (‘yes’ or ‘no’) since 2013: (i) eight of which apply only to households with adults only (19 years old or more); and (ii) six items apply to households with children and/or adolescents^(25)^.

The questions were answered by the reference person in the family responsible for the purchasing and preparation of meals. In addition, the sample design of both surveys (PNAD and POF) considered the country’s master sampling framework, allowing for valid comparison of the FI trends across time in the country.

### Other study variables

The analyses included information about the location of the household (urban/rural area), the region of the country (North, Northeast, Midwest, Southeast and South), along with household socio demographic characteristics such as the number of residents (up to 3, 4 to 6, 7 or more), gender (male/female) and race/skin color (white, black or mulattos) of reference person according to the classification adopted by the IBGE^(35)^. The age groups of household members (0-4 years, 5-17 years, 18-49 years, 50-64 years, and 65 years or more) and the household per capita income quintile were also evaluated.

### Data analysis

Point prevalence and corresponding 95% confidence intervals of households and residents were estimated by FS/FI strata. Contrasts in extreme groups *g*_*1*_*=*FS and *g*_*2*_*=*severe FI explored variations over the periods Δ_1_ =2004-2013 and Δ_2_ =2013-2018. Accordingly, the contrasts *C* per FS/FI strata *g=1,2* and periods D*=1,2* are given by[*C*_Δ_ = (*p*_*y* 2_ − *p*_*y*1_)/*p*_*y*1_]_*g*_, where *p*_*y2*_ and *p*_*y1*_ respectively stand for the proportions (prevalence) in years 2013 and 2004 when Δ_1_; and in years 2018 and 2013 when Δ_2_. Secular trends were plotted by survey year. Analyses using the ‘*svy*’ command of Stata 16 accounted for the complex sample design^(36)^.

## RESULTS

Figure 1 shows the secular trend of the prevalence of FS/FI according to the four household surveys carried out in Brazil. Following a gradual and significant increase in FS between 2004 and 2013, the percentage between 2013 and 2018 reverses to even lower levels then in 2004. Consistent with this finding, severe FI declined substantially between 2004 and 2013, but showed an upturn from 2013-2018.

**Fig.1.**
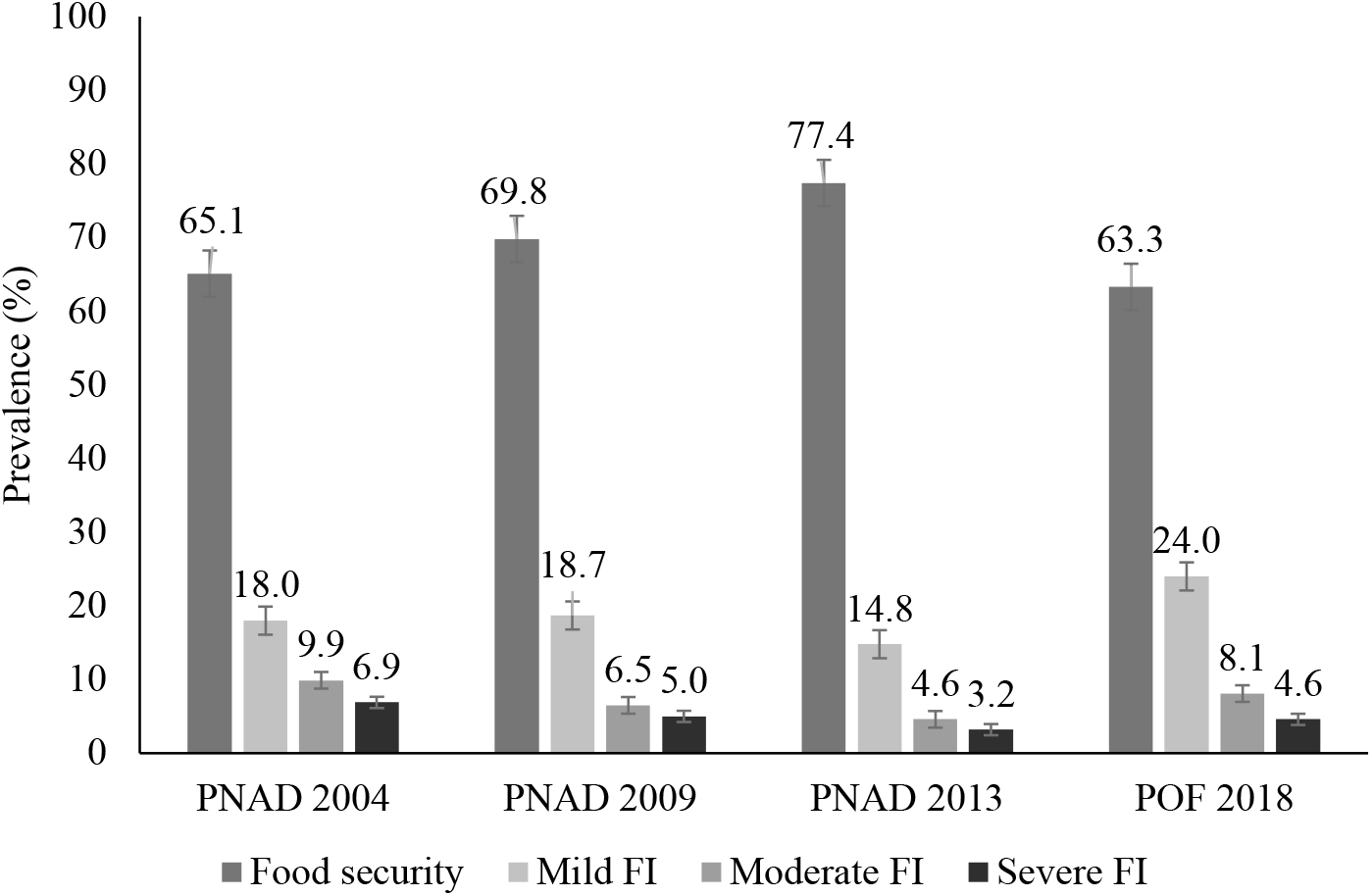
Prevalence (%) of food security and food insecurity (FI) levels. Brazil, 2004-2018.

In 2013, almost 51.5 million households had regular and permanent access to sufficient quality food, without having to compromise access to other essential needs such as housing and health care. By 2018, the POF showed that this figure had dropped to approximately 43.5 million households. The number of households experiencing moderate FI almost doubled in the 2013-2018 period (2.9 and 5.6 million, respectively), with more than one million new households experiencing severe FI in 2018 than in 2013. This pattern was found in all regions of the country, with the greatest difference seen in the Midwest region. In absolute terms, in 2018 the North and Northeast regions continued to present the highest proportion of households in FI, while the Southeast and South regions of Brazil had the highest rates of FS (Supplement 1).

Figure 2 presents secular trends in severe FI according to demographic and socioeconomic variables. The increase in severe FI between 2013 and 2018 occurred in both rural and urban areas (Fig. 2a). Notably, the North and Midwest regions showed the highest percentage increases in severe FI when compared to other regions of the country (Fig. 2b). The highest percentage increases in severe FI occurred in households with 7 or more residents (Fig. 2c), those with women as reference (Fig. 2d), and those comprised of blacks or mulattos (Fig. 2e). The presence of residents above 5 and less than 49 years of age increased the risk of severe household FI (Fig. 2f).

**Fig. 2.**
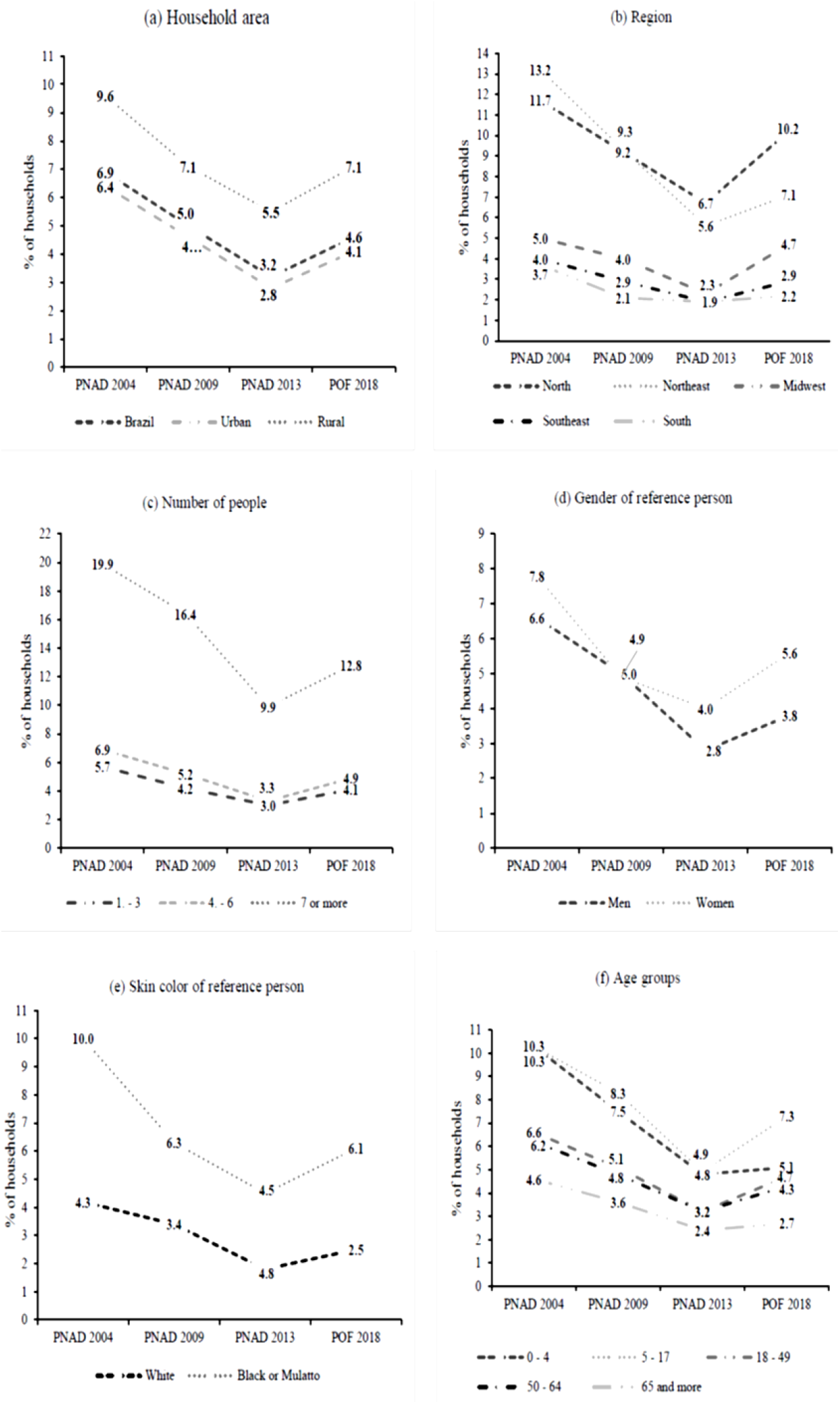
Secular trends in severe food insecurity prevalence according to household characteristics. Brazil, 2004-2018.

Table 1 shows the percentage changes in the extreme FS and severe FI strata for the 2004-2013 and 2013-2018 periods. Regarding FS, there was an increase of 18.9% in the first period, followed by a decrease of 18.2% in the second. The same pattern occurred in both urban and rural areas. The Northeast region held the greatest increase in FS during the first period (+ 33.4%). See Supplement 2 for details.

**Table 1.** Prevalence variation (Δ%) of food security and severe food insecurity (FI) among Brazilian surveys.Brazil, 2004-2018.

| Sociodemographic variables | $\Delta\%$ Food Security* | | $\Delta\%$ Severe FI* | |
| --- | --- | --- | --- | --- |
| | $\Delta_1^\dagger$<br>(2004 and 2013) | $\Delta_2^\dagger$<br>(2013 and 2018) | $\Delta_1^\dagger$<br>(2004 and 2013) | $\Delta_2^\dagger$<br>(2013 and 2018) |
| *Household area* |  |  |  |  |
| Brazil | +18.9 | -18.2 | -53.6 | +43.8 |
| Urban | +19.2 | -18.4 | -56.3 | +46.4 |
| Rural | +15.1 | -17.2 | -42.7 | +29.1 |
| *Region* |  |  |  |  |
| North | +19.4 | -32.7 | -43.7 | +52.2 |
| Northeast | +33.4 | -19.7 | -57.6 | +26.8 |
| Midwest | +18.7 | -20.8 | -54.0 | +104.3 |
| Southeast | +17.3 | -19.5 | -52.5 | +52.6 |
| South | +11.2 | -6.8 | -48.6 | +15.8 |
| *Household characteristics* |  |  |  |  |
| *Number of residents* |  |  |  |  |
| Until 3 | +12.4 | -16.9 | -47.4 | +26.7 |
| 4;6 | +20.3 | -24.2 | -52.2 | +54.5 |
| 7 or more | +40.8 | -35.7 | -50.3 | +30.3 |
| *Gender of reference person* |  |  |  |  |
| Man | +19.9 | -15.2 | -57.6 | +35.7 |
| Women | +16.9 | -21.1 | -48.7 | +40.0 |
| *Skin color from reference person* |  |  |  |  |
| White | +15.4 | -12.9 | -58.1 | +38.9 |
| Black or Mulatto | +27.6 | -22.4 | -55.0 | +35.6 |
| Others | -4.7 | +0.1 | +5.0 | +9.5 |
| *Age groups (years)* |  |  |  |  |
| 0;4 | +32.2 | -24.0 | -53.4 | +6.3 |
| 5;17 | +29.7 | -26.0 | -52.4 | +49.0 |
| 18;49 | +20.2 | -22.2 | -51.5 | +46.9 |
| 50;64 | +15.9 | -17.0 | -49.2 | +34.4 |
| 65 or more | +14.9 | -11.8 | -47.8 | +12.5 |
| *Family per capita income (quintiles)* |  |  |  |  |
| 1<sup>st</sup> | +87.6 | -45.3 | -54.0 | +48.5 |
| 2<sup>nd</sup> | +39.0 | -35.3 | -54.9 | +73.0 |
| 3<sup>rd</sup> | +15.9 | -30.8 | -46.7 | +62.5 |
| 4<sup>th</sup> | +7.7 | -22.4 | -47.4 | +120.0 |
| 5<sup>th</sup> | +1.7 | -10.5 | -40.0 | +200.0 |

\*Estimated by the Brazilian Household Food Insecurity Measurement Scale (*Escala Brasileira de Insegurança Alimentar, EBIA*) applied in Brazilian National Households Sample Surveys (*Pesquisas Nacionais por Amostras de Domicílios [PNADs]*) from 2004 to 2013 (IBGE, 2006; IBGE, 2010; IBGE 2013) and in Household Budget Survey 2017/2018 (*Pesquisa de Orçamentos Familiares* (IBGE, 2020); †Variation (C) considering the proportions of FS/FI strata $g=1,2$ and periods
$D=1,2$ are given by $\left[ C_{\Delta} = \left( p_{y2} - p_{y1} \right) / p_{y1} \right]_g$ , where $p_{y2}$ and $p_{y1}$ respectively stand for the proportions (prevalence) in years 2013 and 2004 when $\Delta_1$ ; and in years 2018 and 2013 when $\Delta_2$ .

The North region stood out as having the largest reduction in severe FI from 2004 to 2013 (–32.7%). There was an increase in FS in the first period followed by a decrease in the second. Households with 7 or more residents had the highest percentage of increase in FS during the first period and the highest percentage of reduction in the following period (+40.8% and –35.7%, respectively). The highest percentages of improvements in FS in the first period were observed in households falling between the 1^st^ and 2^nd^ quintiles of family income per capita; they were the same quintiles with the highest percentages of worsening in the following period (Table 1).

Consistent with the FS trends, a significant reduction in severe FI in the 1st period (–53.6%) was followed by an increase in the 2nd period (+43.8%). Among households located in rural areas, changes in severe FI over time were less pronounced than the national average (–42.7% and +29.1% in the 1st and 2nd periods, respectively). The reduction in severe FI in the 1st period was greater in the Northeast (–57.6%) and Midwest (–54%) regions of the country, while in the 2nd period, the highest percentages increases in severe FI were for the Midwest (+104.3%), Southeast (+52.6%) and North (+52.2%) regions. The significant reduction of severe FI in the 1st period was stronger in households with 4 to 6 residents (–52.2%), whose reference person was male (–57.6%) and of white skin color (–58.1%), with children under 4 years old (–53.4%) and in the lowest quintiles of family income per capita. In the 2nd period, the lower increases in severe FI were observed in households with children under 4 years old (+6.3%) and with members over 65 years old (+12.5%) (Table 1).

## DISCUSSION

Since the validation of the EBIA, the measurement of household FI in the PNADs (2004, 2009 and 2013) has contributed to the discussion on the direction the country should take when planning programs and initiatives to guarantee FNS for the Brazilian population. The result of these actions was reflected in the strong reduction in FI, mainly in its most severe form in the first period of almost 10 years^(33)^.

The analyses of this study, which are based on adding the POF 2018 data, revealed a setback in Brazil regarding the human right to food in the context of increasing social inequalities. Following a downward trend for a decade since 2004, FS decreases from 2013 onwards; within a five-year period, by 2018, an increase in all forms of FI is quite poignant. ^(29)^. This may be explained by major disruptions in access to healthy foods and in some instances even to enough amount of food, regardless of quality.

In addition, regional FI inequities persisted between 2004 and 2013^(32,33)^ and worsened by 2018, erasing the previous advances made in the country^(29)^. It is disquieting that less than half of the North and Northeast regions’ residents had full and regular access to food. Households in the northern region of the country, which include the states of the Amazon rain forest that is home to the majority of the traditional indigenous people of Brazil, an also of slave-descendant communities known as *Quilombolas*, and other traditional or native populations. Households in these regions were found to be four times more likely to experience severe FI than households in the more developed regions of the country (South and Southeast regions). The prevalence of severe FI in the Northeast – an historically, socioeconomically, and environmentally vulnerable semi-arid region – was three times that of other regions. However, this does not imply that there were no families living with hunger in the better off regions of Brazil. According to the 2018 data, in the South and Southeast regions, more than 1.3 million households, corresponding to approximately 5 million people, endured severely restricted access to food^(37)^.

The sociodemographic relationship with FI were similar across both time periods examined^(14)^. Severe FI was consistently higher in households with a more density of residents, with low income, and whose reference person was female and declared herself as being black/mulatto^(38)^. Households with at least one of these conditions experienced a strong reduction in severe FI in the period between 2004 and 2013, followed by an increase between 2013 and 2018, maintaining the strong inequities previously documented^(39)^.

The risk components reflecting strong inequities in FI in Brazil were also documented for some household demographics including the presence in the household of members of different age group distribution^(40)^. Interestingly, despite the high prevalence of severe FI in households with at least one child under 5 years old, this group experienced the smallest increase between 2013 and 2018. In turn, households with at least one older adult (>65) consistently had higher prevalence of FS and a lower prevalence of severe FI. It is possible that cash transfer programs have helped buffer children from experiencing severe FI, at least to some extent. Palmeira et al.^(40)^ endorsed this hypothesis in an area of extreme climatic and social vulnerability in the Northeast. Extremely poor families who were beneficiaries of the Brazilian conditional cash transfer called ‘*Programa Bolsa Família – PBF*’ and who had small children receive a slightly higher financial benefit. As the PBF has good coverage and targeting^(41)^, the beneficiary families would be expected to be more protected from having severe FI. In turn, older adults with insufficient incomes are entitled to a continuous benefit of a minimum wage per month (Benefit of Continuous Installment, in Portuguese ‘*Benefício de Prestação Continuada – BPC’*), which has also contributed to poverty reduction^(42)^. In this way, poor families with older adult BPC beneficiaries would be expected to also be more protected from FI.

Our findings suggest that keeping social protection programs such as those described above, even in the context of fiscal austerity, may help protecting households against FI in the most vulnerable groups. This is consistent with findings from other parts of the world^(6,9)^.

In the period from 2004 to 2013, favorable economic and political conditions in Brazil, along with public policies to promote the FNS. This enabled not only greater access of the Brazilian population to food but also protected the most vulnerable populations, including residents in the North and Northeast regions, children under 5 years of age, older adults and the lowest income households^(43)^. IBGE data comparing socioeconomic and demographic indicators derived from the 2000 and 2010 Brazilian Demographic Census confirmed that Brazilian society underwent important improvements during the past decade, which was reflected in significant FS improvements^(44)^.

Between 2004 and 2013, there was indeed a real (deflated) increase in the on the order of 30% for average incomes in the employed population. This positive economic outcome partly reflected an increase in the real value of Brazilian minimum wage by more than 60%, coupled with the reduction of open unemployment over this period of time^(45)^. In the same period, the Brazilian cash conditional transfer PBF was created, and its coverage was gradually expanded, reaching 13.8 million families in 2013^(46)^. Households engaged in family farming and in a situation of food vulnerability, received relevant incentives through a program designed for the early acquisition of their products (Food Acquisition Program, in Portuguese *Programa de Aquisição de Alimentos*), construction of water cisterns in the regions of the Brazilian semiarid region, used for both consumption and food production, and the expansion of financing programs for famers’ production, including the PRONAF^(47)^. It is also relevant that in this period there was a strong governance model based on the national, state, and municipal councils for food and nutritional security (CONSEAs), which had a strong participation of civil society^(9)^.

The effect of social changes observed in Brazil on reducing FI in some municipalities corroborates these study^(48-50)^. In their study, Palmeira et al.^(8)^ observed a reduction in FI followed by improvement in socio-economic indicators in two waves of data collected in representative samples in a poor municipality in Rio de Janeiro (Southeast Brazil). Their findings support the hypothesis that these changes occurred due to improvements in socioeconomic indicators and in participation in the PBF.

The improvements in FS observed between 2004 and 2013 may also be explained by specific government policy initiatives deployed during this period. Both structural and emergency measures, resulted in households’ increased ability to access food, but above all, they led to reduced poverty and extreme poverty in Brazil^(41)^. In 2003, there were about 42 million Brazilians below the poverty line and almost 13 million in extreme poverty^(51)^. By 2014, poverty had been reduced by one-third (14 million), and extreme poverty was reduced by more than half, to approximately 5 million. The FAO attributed these advances in Brazil to the government’s commitment to more equitable public policies^(52)^.

One limitation of this study is that Brazilian surveys including FS/FI indicators have been scheduled to take place only at 5-year intervals, which precludes a more refined view of what happened between 2014 and 2017. However, indirect indicators and FNS predictors have already informed the food access crisis, which was confirmed with the 2018 findings. An example is the progressive increase in poverty and extreme poverty, and in unemployment from 2014 to 2017. According to the Institute for Applied Economic Research (*Instituto de Pesquisa Econômica e Aplicada - IPEA*), the percentage of households experiencing extreme poverty increased by 78% ^(46)^, and the proportion of unemployment doubled in the same period of time^(53)^. Although there is a lack of information on FS/FI for the period of great social and political crisis in Brazil that began in 2015 and worsened in 2016, it was anticipated that FI would have increase as poverty and unemployment are two of their major determinants ^(22)^.

Looking ahead, it may well be that the increases in FI and hunger have already worsen in the current Brazilian context as a result of an additional crisis. While up to 2018 we only had to deal with the effect of the political and economic crises on FNS, a major health-related one has been added as of the beginning of 2020 with COVID – 19 pandemic^(54)^. A study carried out by the United Nations International Children’s Emergency Fund (UNICEF) and published in July 2020 ^(55)^ evaluating the primary and secondary impacts of the COVID-19 pandemic on Brazilian children and adolescents revealed an extremely serious state of affairs concerning FS/FI. Accordingly, whereas 64% of adults above the age of 18 were working before the pandemic, this percentage dropped to 50% within end of Feb to July 2020. Furthermore, the pandemic caused a drop in income for more than half of the adult Brazilian population, and further aggravated in the households holding children and adolescents lived ^(54)^.

## CONCLUSIONS

This study showed a conspicuous regression in the advances achieved in the 2004-2013 period regarding the reduction of FI and hunger in Brazil, and the mitigation of existing inequalities. These data make it clear that Brazil will not be able to reach the SDGs with regards to FI and hunger reductions and this is in turn will strongly prevent Brazil from meeting the rest of the SDGs^(1)^. The second point that this article highlighted is the increase in regional, gender, and racial inequalities for severe FI, which definitively accentuated in the period between 2013 and 2018. It is likely that if the economic recession and reduced spending on FNS policies that started since 2015 in Brazil continues, the FI situation in Brazilian households will continue to deteriorate. Not addressing this reality could strongly and negatively affect the future development of Brazil and the wellbeing of all its people.

## Data Availability

This paper used secondary data available to the public domain by Brazilian Institute of Geography and Statistics (IBGE).

## Ethical Standards Disclosure

In according to Resolution No. 466, of December 12, 2012 from National Committee of Ethics in Research (CONEP), researches that used secondary data available to the public domain, as occurs in this research that used data available to the public domain by Brazilian Institute of Geography and Statistics (IBGE), the approval by an local Ethic Committee CEP-CONEP System was not required.

---

**Supplement 1.**
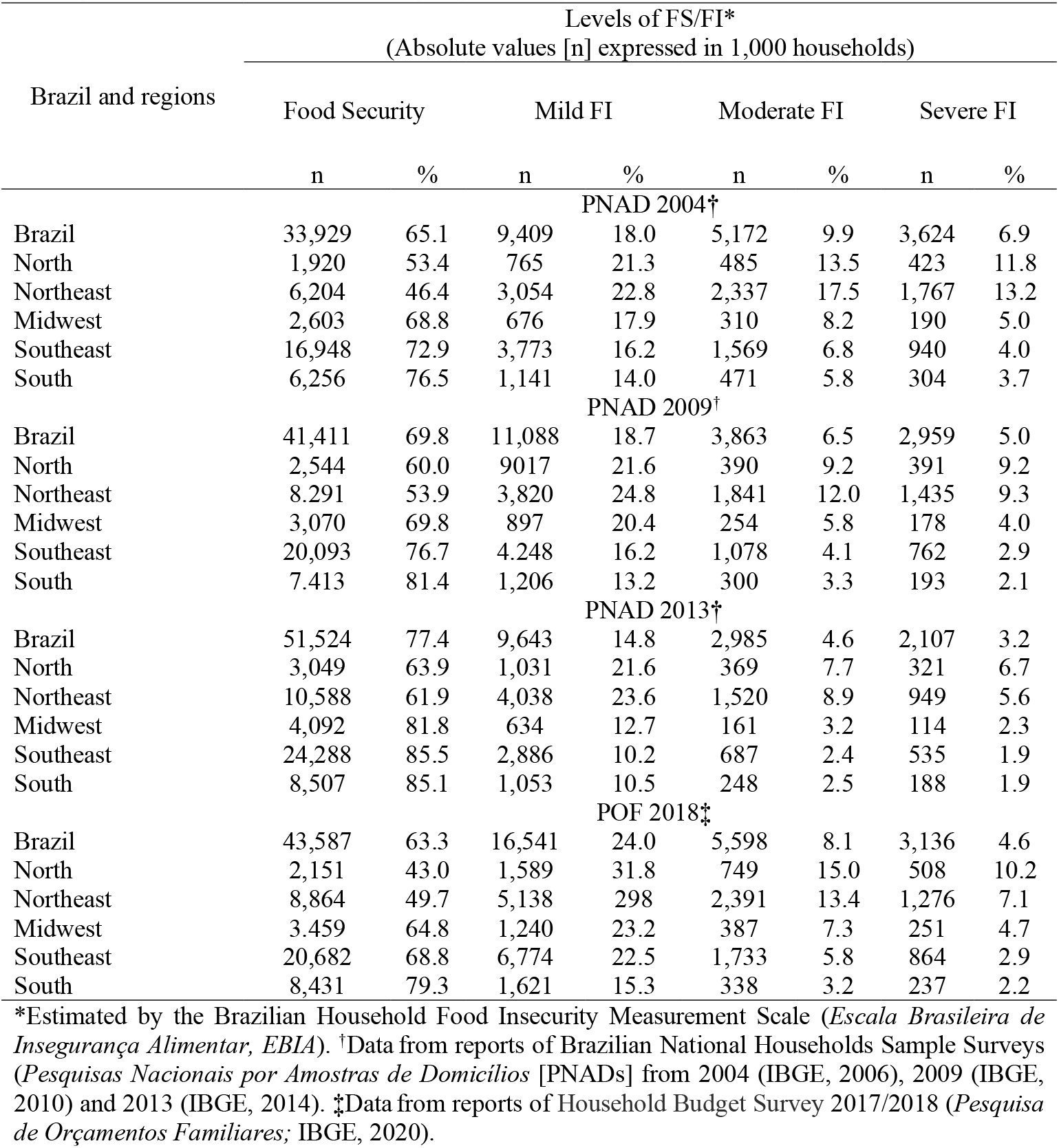
Absolutes and percentage values (%) from Brazilian households in according to the food security/food insecurity (FS/FI) levels by regions and year of evaluation. Brazil, 2004-2018.

| Brazil and regions | Levels of FS/FI*<br>(Absolute values [n] expressed in 1,000 households) |  |  |  |  |  |  |  |
| --- | --- | --- | --- | --- | --- | --- | --- | --- |
|  | Food Security |  | Mild FI |  | Moderate FI |  | Severe FI |  |
|  | n | % | n | % | n | % | n | % |
| PNAD 2004† |  |  |  |  |  |  |  |  |
| Brazil | 33,929 | 65.1 | 9,409 | 18.0 | 5,172 | 9.9 | 3,624 | 6.9 |
| North | 1,920 | 53.4 | 765 | 21.3 | 485 | 13.5 | 423 | 11.8 |
| Northeast | 6,204 | 46.4 | 3,054 | 22.8 | 2,337 | 17.5 | 1,767 | 13.2 |
| Midwest | 2,603 | 68.8 | 676 | 17.9 | 310 | 8.2 | 190 | 5.0 |
| Southeast | 16,948 | 72.9 | 3,773 | 16.2 | 1,569 | 6.8 | 940 | 4.0 |
| South | 6,256 | 76.5 | 1,141 | 14.0 | 471 | 5.8 | 304 | 3.7 |
| PNAD 2009† |  |  |  |  |  |  |  |  |
| Brazil | 41,411 | 69.8 | 11,088 | 18.7 | 3,863 | 6.5 | 2,959 | 5.0 |
| North | 2,544 | 60.0 | 9017 | 21.6 | 390 | 9.2 | 391 | 9.2 |
| Northeast | 8,291 | 53.9 | 3,820 | 24.8 | 1,841 | 12.0 | 1,435 | 9.3 |
| Midwest | 3,070 | 69.8 | 897 | 20.4 | 254 | 5.8 | 178 | 4.0 |
| Southeast | 20,093 | 76.7 | 4,248 | 16.2 | 1,078 | 4.1 | 762 | 2.9 |
| South | 7,413 | 81.4 | 1,206 | 13.2 | 300 | 3.3 | 193 | 2.1 |
| PNAD 2013† |  |  |  |  |  |  |  |  |
| Brazil | 51,524 | 77.4 | 9,643 | 14.8 | 2,985 | 4.6 | 2,107 | 3.2 |
| North | 3,049 | 63.9 | 1,031 | 21.6 | 369 | 7.7 | 321 | 6.7 |
| Northeast | 10,588 | 61.9 | 4,038 | 23.6 | 1,520 | 8.9 | 949 | 5.6 |
| Midwest | 4,092 | 81.8 | 634 | 12.7 | 161 | 3.2 | 114 | 2.3 |
| Southeast | 24,288 | 85.5 | 2,886 | 10.2 | 687 | 2.4 | 535 | 1.9 |
| South | 8,507 | 85.1 | 1,053 | 10.5 | 248 | 2.5 | 188 | 1.9 |
| POF 2018‡ |  |  |  |  |  |  |  |  |
| Brazil | 43,587 | 63.3 | 16,541 | 24.0 | 5,598 | 8.1 | 3,136 | 4.6 |
| North | 2,151 | 43.0 | 1,589 | 31.8 | 749 | 15.0 | 508 | 10.2 |
| Northeast | 8,864 | 49.7 | 5,138 | 29.8 | 2,391 | 13.4 | 1,276 | 7.1 |
| Midwest | 3,459 | 64.8 | 1,240 | 23.2 | 387 | 7.3 | 251 | 4.7 |
| Southeast | 20,682 | 68.8 | 6,774 | 22.5 | 1,733 | 5.8 | 864 | 2.9 |
| South | 8,431 | 79.3 | 1,621 | 15.3 | 338 | 3.2 | 237 | 2.2 |

\*Estimated by the Brazilian Household Food Insecurity Measurement Scale (*Escala Brasileira de Insegurança Alimentar, EBLA*). †Data from reports of Brazilian National Households Sample Surveys (*Pesquisas Nacionais por Amostras de Domicílios* [PNADs] from 2004 (IBGE, 2006), 2009 (IBGE, 2010) and 2013 (IBGE, 2014). ‡Data from reports of Household Budget Survey 2017/2018 (*Pesquisa de Orçamentos Familiares*; IBGE, 2020).

**Supplement 2.** Percentual of food security and severe food insecurity (FI) in Brazil according to sociodemographic variables. Brazil 2004-2018.

| Sociodemographics variables | Food security* |  |  |  | Severe FI* |  |  |  |
| --- | --- | --- | --- | --- | --- | --- | --- | --- |
|  | PNAD 2004<sup>†</sup> | PNAD 2009<sup>†</sup> | PNAD 2013<sup>†</sup> | POF 2018<sup>‡</sup> | PNAD 2004<sup>†</sup> | PNAD 2009<sup>†</sup> | PNAD 2013<sup>†</sup> | POF 2018<sup>‡</sup> |
| *Household area* |  |  |  |  |  |  |  |  |
| Brazil | 65.1 (64.4;65.7) | 69.8 (69.2;70.42) | 77.4 (76.5;77.6) | 63.3 (62.53;64.08) | 6.9 (6.6;7.2) | 5.0 (4.7;5.2) | 3.2 (3.1;3.4) | 4.6(4.3;4.8) |
| Urbano | 66.7 (66.0;67.3) | 70.7 (70.0;71.3) | 79.5 (78.6;79.6) | 64.9 (64.6;71.0) | 6.4(6.2;6.7) | 4.6 (4.7;5.2) | 2.8(2.7;3.0) | 4.1 (3.8;4.4) |
| Rural | 56.2 (54.0;58.4) | 64.8 (62.8;66.8) | 64.7 (62.7;66.0) | 53.6 (51.9;55.2) | 9.6 (8.4;10.9) | 7.1 (6.3;8.0) | 5.5 (5.0;6.1) | 7.1 (6.4;7.9) |
| *Region* |  |  |  |  |  |  |  |  |
| North | 53.5 (51.0;55.9) | 60.0 (58.0;61.9) | 63.9 (61.6;65.1) | 43.0 (40.9;45.1) | 11.7(10.3;13.3) | 9.2 (8.1;10.4) | 6.7 (6.1;7.4) | 10.2(9.0;11.3) |
| Northeast | 46.4 (44.6;48.2) | 53.9 (52.3;55.4) | 61.9 (60.3;61.7) | 49.7 (48.5;50.8) | 13.2(12.1;14.3) | 9.3 (8.6;10.0) | 5.6 (5.2;6.1) | 7.1 (6.6;7.6) |
| Midwest | 68.9 (67.0;70.6) | 69.8 (68.0;71.5) | 81.8 (80.1;83.0) | 64.8 (62.4;66.8) | 5.0 (4.5;5.5) | 4.0(3.6;4.4) | 2.3 (2.0;2.7) | 4.7 (4.0;5.5) |
| Southeast | 72.9 (72.0;73.8) | 76.7 (75.8;77.6) | 85.5 (84.5;85.9) | 68.8 (67.4;70.2) | 4.0 (3.7;4.3) | 2.9 (2.7;3.1) | 1.9 (1.7;2.1) | 2.9 (2.4;3.2) |
| South | 76.5 (75.2;77.8) | 81.3 (80.4;82.2) | 85.1 (84.5;86.9) | 79.3 (77.9;80.7) | 3.7 (3.3;4.1) | 2.1 (1.9;2.3) | 1.9 (1.6;2.2) | 2.2 (1.8;2.6) |
| *Household characteristics* |  |  |  |  |  |  |  |  |
| *Number of residents* |  |  |  |  |  |  |  |  |
| Until 3 | 71.8 (71.2;72.5) | 74.8 (74.1;75.3) | 80.7 (80.2;81.2) | 67.1 (66.2;67.9) | 5.7 (5.4;5.9) | 4.2 (3.9;4.4) | 3.0 (2.8;3.1) | 3.8 (3.5;4.0) |
| 4;6 | 60.1 (59.3;60.9) | 64.4 (63.7;43.3) | 72.4 (71.5;73.0) | 54.0 (52.8;55.2) | 6.9 (6.5;7.3) | 5.2 (4.8;5.4) | 3.3 (3.1;3.6) | 5.1 (4.6;5.6) |
| 7 or more | 34.8 (33.1;36.5) | 41.2 (39.7;43.3) | 49.0 (47.2;51.6) | 31.5 (28.1;35.1) | 19.9 (18.4;21.5) | 16.5 (15.0;18.1) | 9.9 8.7;11.3) | 12.9(11.1;15.2) |
| *Gender of reference person* |  |  |  |  |  |  |  |  |
| Man | 65.7 (64.9;66.4) | 69.7 (69.20;70.3) | 78.8(78.2;79.3) | 66.8 (65.8;67.7) | 6.6 (6.2;7.0) | 5.0(4.7;5.2) | 2.8 (2.6;3.0) | 3.8 (3.5;4.0) |
| Women | 63.4 (62.5;64.2) | 69.9 (69.2;70.6) | 74.1 (73.9;74.7) | 58.5 (57.4;59.5) | 7.8 (7.4;8.2) | 4.9 (4.6;5.2) | 4.0 (3.7;4.2) | 5.6 (5.2;6.0) |
| *Skin color of reference person* |  |  |  |  |  |  |  |  |
| White | 73.5 (72.9;74.2) | 75.6 (75.0;76.2) | 84.8 (84.3;85.2) | 73.9 (73.0;74.9) | 4.2 (4.0;4.5) | 3.4 (3.2;3.6) | 1.8 (1.7;2.0) | 2.5 (2.3;2.8) |
| Black or Mulatto | 55.0 (54.1;55.9) | 64.6 (63.9;65.4) | 70.2 (69.5;70.8) | 54.5 (53.5;55.4) | 10.0 (9.5;10.6) | 6.3 (6.0;6.6) | 4.5 (4.3;4.8) | 6.1 (5.8;6.5) |
| Others | 75.8 (71.7;79.4) | 73.6 (70.2;76.8) | 77.2 (76.5;77.6) | 72.3 (67.1;76.9) | 4.0 (2.6;6.4) | 4.6(3.3;6.4) | 4.2 (2.8;6.2) | 4.6 (3.2;6.6) |
| *Age groups (years)* |  |  |  |  |  |  |  |  |
| 0;4 | 49.4 (48.8;49.9) | 56.6 (55.9;57.2) | 65.8 (64.8;66.1) | 50.0 (48.4;51.5) | 10.3 (10.0;18.9) | 7.5(7.1;7.8) | 4.8 (4.6;5.2) | 5.1 (4.5;5.7) |
| 5;17 | 51.5 (51.2;51.9) | 57.9 (57.5;58.2) | 66.8 (66.0;66.8) | 49.4 (48.2;50.6) | 10.3 (10.1;10.5) | 8.3 (8.1;8.5) | 4.9 (4.8;5.2) | 7.3 (6.6;7.9) |
| 18;49 | 62.9 (62.6;63.1) | 67.4 (67.2;67.6) | 75.6 (75.0;77.4) | 58.8 (57.9;59.7) | 6.6 (6.5;6.7) | 5.1(4.9;5.2) | 3.2 (3.1;3.3) | 4.7 (4.4;5.0) |
| 50;64 | 67.9 (67.4;68.4) | 72.1 (71.7;72.6) | 78.7 (78.1;78.8) | 65.3 (64.2;66.4) | 6.2(6.0;6.5) | 4.8 (4.5;4.9) | 3.2 (3.1;3.4) | 4.3 (4.0;4.7) |
| 65 or more | 71.7 (71.1;72.3) | 76.2 (75.6;76.7) | 82.4 (81.5;82.4) | 72.7 (71.5;73.9) | 4.6 (4.3;4.9) | 3.6 (3.3;3.7) | 2.4 (2.3;2.6) | 2.7 (2.4;3.0) |
| *Family per capita income (quintiles)* |  |  |  |  |  |  |  |  |
| 1<sup>st</sup> | 26.6 (25.3;27.9) | 39.4 (38.1;40.7) | 49.9 (48.5;51.1) | 27.3 (25.9;28.9) | 21.1 (19.9;22.2) | 15.0 (14.2;15.8) | 9.7 9.1;10.3) | 14.4 (13.2;15.6) |
| 2<sup>nd</sup> | 49.5 (48.4;50.6) | 59.4 (58.4;60.4) | 68.8 (67.9;69.7) | 44.5 (42.8;46.2) | 8.2 (7.7;8.7) | 5.8(5.4;6.2) | 3.7 (3.4;4.1) | 6.4 (5.7;7.2) |
| 3<sup>rd</sup> | 68.7 (67.8;69.6) | 72.2 (71.4;73.1) | 79.6 (78.8;80.3) | 55.1 (53.5;56.6) | 4.5 (4.2;4.8) | 3.4 (43.2;3.7) | 2.4 (2.2;2.7) | 3.9 (3.5;4.6) |
| 4<sup>th</sup> | 81.6 (80.8;82.2) | 81.7 (81.1;82.4) | 87.9 (87.3;88.5) | 68.2 (67.2;69.9) | 1.9(1.7;2.1) | 1.5 (1.3;1.7) | 1.0 (0.8;1.2) | 2.2 (1.9;2.6) |
| 5<sup>th</sup> | 93.4 (93.5;94.3) | 91.7 (91.2;92.2) | 95.0 (94.6;95.4) | 85.0 (84.1;86.0) | 0.5(0.4;0.6) | 0.4(0.3;0.5) | 0.3 (0.2;0.4) | 0.8 (0.6;0.9) |

\*Food security/food insecurity levels estimated by the Brazilian Household Food Insecurity Measurement Scale (*Escala Brasileira de Insegurança Alimentar, EBI4*); <sup>†</sup>Estimated using the microdata of the Brazilian National Households Sample Surveys (*Pesquisas Nacionais por Amostras de Domicílios* [PNADs] from 2004 (IBGE, 2006), 2009 (IBGE, 2010) and 2013 (IBGE, 2014); <sup>‡</sup>Estimated using the microdata of the Household Budget Survey 2017/2018 (*Pesquisa de Orçamentos Familiares*; IBGE, 2020).

